# COVID-19 vaccine attitude and its predictors among people living with chronic health conditions in Ibadan, Nigeria

**DOI:** 10.1101/2022.01.27.22269947

**Authors:** Lucia Yetunde Ojewale, Rotimi Felix Afolabi, Adesola Ogunniyi

**Author notes:** Corresponding author (LYO).

## Abstract

**Background:** Globally COVID-19 has caused death among millions of people and new cases continue to be reported daily, including in Nigeria. With the efforts of the Nigerian government to ensure everyone gets vaccinated, the vaccination attitude and its predictors among persons with chronic health conditions remains unclear. The study was therefore conducted to assess vaccination attitude and determine its associated factors among people living with chronic health conditions.

**Methods:** A descriptive cross-sectional study was conducted among 423 patients attending the medical outpatient clinic of University College Hospital, Ibadan, Oyo State, Nigeria; before COVID-19 vaccination commencement. Data were collected on socio-demographic and COVID-19 related characteristics, via Open Data Kit (ODK) software. The Vaccine Attitude Examination (Vax) Scale including its four subscales was adopted to assess attitude towards COVID-19 vaccine uptake. The main outcome was vaccine attitude status defined as positive if a VAX sum score was above the median value; otherwise, non-positive’. Data were analysed using Chi-square and multivariate logistic regression analyses at 5% significance level.

**Results:** Hypertension (27.4%), diabetes mellitus (22.0%) and heart conditions/diseases (19.6%) were the top three conditions being managed by the participants. The overall proportion of patients with a positive attitude towards the uptake of COVID-19 vaccination was 46.6%; while 29.6% trusted the vaccine benefit, 46.6% were not worried about the aftermath effect of the vaccine and 11.1% were not concerned about the vaccine commercial profiteering. Factors associated with overall vaccine attitude were level of education, income, knowledge of COVID-19, living room arrangement, and confidence in government (p<0.05). The main influential factor on general vaccine positive attitude and the four subscales was confidence in the government.

**Conclusion:** Less than half of people living with a chronic medical condition had a positive attitude towards the COVID vaccine. The attitudes are mediated strongly by confidence in the government and several sociodemographic and COVID related characteristics. A lot still needs to be done to achieve the prescribed herd immunity.

## Introduction

The impact of the COVID-19 pandemic on social, economic and political life is unprecedented. Lockdown was effected in several countries to guarantee limited contact between individuals and to ensure that citizens observed social distancing, which admissibly had helped curtail virus transmission [1]. However, a huge economic price was paid as many countries suffered economic losses [2]. This led to easing the lockdown which soon resulted in an increase in COVID 19 infection and mortality[3]. The health system has not been able to effectively cater for the needs of those suffering from Acute Respiratory Distress (ARD) and SARS COVID– 2 pneumonias, most especially in Africa [4]. These negative effects led to efforts to prevent further transmission through the development of vaccines. Vaccine development is seen as crucial to ending the pandemic, [5]. As of late February 2021, COVID-19 vaccines were already available and were being administered to people mostly in high-income countries like the United Kingdom, United States, Canada, China, among others,[6]. In line with this and the effort by the World Health Organisation to ensure that the vaccines get to low- and middle-income countries, there is a need to ensure that it is well received by the general population, particularly those with chronic conditions.

Nonetheless, vaccine availability does not necessarily translate to the uptake. It has been suggested that apart from prioritizing vaccine administration, other important factors that would affect vaccine distribution include the capability of the health system in ensuring that the vaccines are made available for people at high risk and the willingness of the people to be vaccinated, [7]. Despite the fatality of COVID-19 and the purported success in developing vaccines, a sceptical attitude continues to trail vaccination in many countries of the world, including developed countries. This phenomenon, sometimes referred to as ‘vaccine hesitancy, has been reported in many countries largely due to vaccine disinformation [8,9]. Latkin et al. (2021) reported that 40.9% of adults in the US mistrusted the vaccine while 16% of adults in the UK had a high level of mistrust, towards the vaccine, [10]. Also, 69% of adult participants were willing to get vaccinated in a study among over 2000 adults in US [11]. The proportion of people intending to get vaccinated is similar to that reported among about a thousand Hong Kong nurses, [1].

It was opined that the proportion of vaccinated people should be greater than two-thirds to achieve ‘herd’ OR public immunity, [1]. Literature has shown that reasons for unwillingness to vaccinate, mistrust of vaccines and poor vaccine intention include low socioeconomic status, lower education, older age, concerns about the unforeseen side effects, mistrust of Government, and poor adherence to COVID-19 prevention guidelines, [1,5,10].

Addressing the aforementioned factors may promote a positive attitude and high intention to be vaccinated. Besides, COVID-19 vaccine recommendation by health care providers was a positive mediating factor towards its uptake among US adults, [11]. Adequate understanding of clients by health care professionals is thus crucial before they can effectively communicate the need for vaccines [12]. Nurses, physicians and other health care workers can thus effectively improve the attitude and uptake of the vaccine among their clients.

Generally, the population at higher risk of death or complications should be vaccinated first due to inadequate supply of COVID-19 vaccine. Many countries have therefore adopted the WHO vaccination guideline of prioritizing people at higher risk of mortality from the disease including older adults and those with chronic conditions like diabetes and chronic kidney disease [13].

Specifically, adults with chronic condition are more likely to be hospitalized due to COVID-19 infection compared to healthy individuals in a study conducted in the United States, [14]. For example, having diabetes increases the risk of developing COVID-19 as well as increases the risk of dying from COVID-19 complications [15]. Studies in Africa including Nigeria have also corroborated the claim that people who have co-morbidities such as diabetes and hypertension are more likely to suffer fatality from COVID-19 infection, [16].

Apart from the vulnerability of people with chronic conditions being reported, the patients themselves are aware of this fact. In a study conducted among Ethiopian living with diabetes and hypertension. many (79%) participants felt that they were more susceptible to COVID-19 death, yet only 10% were involved in a good level of COVID-19 prevention measures, [17]. The poor attitude towards COVID-19 prevention could be carried over to the reception of the vaccine. Hence, the need to assess vaccination attitude.

Many studies have examined the COVID-19 vaccination attitude but few of such were conducted in Nigeria. The attitude towards vaccination and its associated factors may differ considerably in Nigeria. Moreover, most studies have focused on the general population, whereas there is a need to ascertain the attitude of people with chronic diseases including diabetes, hypertension and chronic kidney disease towards vaccination since they are among the high priority group that need to get vaccinated. Against this background, the study aimed at assessing COVID-19 vaccine attitude among persons living with chronic health conditions, receiving treatment in University College Hospital Ibadan, Nigeria, and to determine its associated factors

## Materials and Methods

### The study design and setting

A cross-sectional study on attitude and intention to COVID-19 vaccine uptake among People Living with Chronic Health Conditions in Ibadan, Nigeria was conducted between March and April 2021. This study is part of a larger study on ‘Covid-19 Vaccine: Attitude, Intention to Vaccinate, Mediating Factors and Interventions towards a Positive Attitudes among People with Chronic Conditions in Ibadan’ The present study was conducted at the Medical Outpatient Clinic of the University College Hospital (UCH), Ibadan, Nigeria. Eligible consenting patients were referred to participate by Counselors after routine patients’ education sessions during which information about the study was provided to all clinic attendees. On average, 20 participants were expected on each clinic day for the two months of data collection.

### Sample size determination and sampling strategy

Based on the assumptions of a 50% prevalence of positive attitude towards the uptake of COVID-19 vaccination among the patients and a 5% desired level of precision, the required minimum sample size was estimated. A total of 423 sample size was estimated for the study after adjusting for a 10% non-response rate. At every clinic visit, eligible consenting participants were selected using a simple random sampling (balloting approach). Daily attendance register at the Record section of the clinic served as the sampling frame. There were 20 “yes” of the total prepared secret ballot-papers, labelled “yes” or “no” for eligible patients who registered on a clinic day. Patient who selected a “yes” was enrolled into the study after written informed consent was obtained, while excluding patient who was very ill and cognitively impaired.

### Data collection

Data collection took place before the vaccination of the general population commenced in Nigeria. Data was collected on socio-demographic variables and COVID-19 related characteristics among the patients by trained research assistants. The interviewers who were postgraduate students in the College of Medicine were trained at a one-day workshop. During the training, they got general orientation about the study objectives, interviewing skills and health research ethics. Each question item was explained as well as how to record the responses. A questionnaire consisting of three (3) sections was used for data collection. The first section consisted of sociodemographic data and predictors of vaccination attitude, based on a literature search. Items included were gender, age, socioeconomic status using the wealth index, [18], employment classification, history of children’s vaccination, daily exposure to news, and self-rated adherence to the COVID-19 guidelines, among others.

The second section was made up of the Vaccine Attitude Examination (Vax) Scale. The VAX scale is an easy to use made up of 12-item questions [19]. The scale consisted of four subscales which provide information on individuals with vaccination resistance. The subscales are: 1) mistrust of vaccine benefit, (2) worries about unforeseen future effects, (3) concerns about commercial profiteering, and (4) preference for natural immunity. A sufficient convergent validity and internal reliability (Cronbach’s alphas = 0.77-0.93) had been established for all four subscales, (19); Wood, Smith, Miller, & O’Carroll, 2019). The scale is rated on a six-point Likert scale (very strongly disagree (coded as 0), …, very strongly agree (coded as 5)). With a maximum possible score of 60, the overall score was dichotomized using the median value as a cut off value

The last section was made up of questions to ascertain contextual influences on COVID-19 vaccine attitude. It was made up of 16 items with three main options: ‘Yes/No/Not sure’

### Data processing and analysis

The analysis started with data cleaning to ensure completeness and consistency. The main outcome variable was a positive attitude towards the uptake of the COVID-19 vaccine. The response to the attitude questions was summed together to generate an attitude score ranging from 0 to 60. Similar scores (ranged 0-15 scores) were generated for each of the four subscales of attitude towards COVID-19 vaccine uptake. Having confirmed the nonnormality of the outcome variable including its subscales’ scores using the Shapiro Wilk normality test (p< 0.05), an overall score above the median value was coded “1” as positive attitude; otherwise, coded “0” as non-positive(20). Independent variables considered were socio-demographics, contextual and COVID-19 related characteristics, see Table 1.

**Table 1.** Definitions of independent variables

| Characteristics | Description |
| --- | --- |
| Age | Patient's age (in years) categorised as: <30, 30-49, 50-64, ≥65 years |
| Sex | Sex of the patient (male or female) |
| Ethnicity | Patient ethnicity (Yoruba, others) |
| Religion | The religion of patients is grouped into two: Christianity, Islam |
| Marital | Patient marital status: Never married, married |
| Employment | Patient employment status: skilled, unskilled |
| Education | Patient highest level of educational attainment categorised as no formal, primary, secondary, higher |
| Income | Patient monthly income (in Naira) categorised as: <30,000, 30,000-49,999, 50,000-99,999, ≥100,000, unknown |
| Assets own | Patient total asset grouped into single (only one asset declared) or multiple (two or more assets declared) |
| Media exposure | Exposure to media is grouped into not exposed and exposed. |
| Cooking fuel | Type of cooking fuel categorised as clean and unclean |
| Drinking water source | Source of drinking water grouped as improved and unimproved |
| Toilet facility | Household toilet facilities: improved; unimproved |
| Waste disposal | Waste disposal practice: hygienic; unhygienic |
| No of rooms | Residential house number of rooms categorised as 1-2, 2-4, >4 |
| No of persons | Number of persons living in a house categorised as 1-2, 2-4, 4-6, >6 |
| Health condition | Patient health challenges grouped as single or multiple diseases |
| COVID-19 knowledge | Rated level of COVID-19 knowledge grouped into poor and good |
| Adherence level | Adherence to the COVID-19 guideline: poor, better |
| Full child vaccination | Participant's children were full vaccination: complete and incomplete/none |
| Conf in govt | Patient rated level of confidence in government in handling pandemic: undecided, low, high |
| Conf in health prof | Patient rated level of confidence in health care professionals: undecided, low, high |

Descriptive statistics such as percentages were used to report the frequency distribution and prevalence of the overall positive attitude towards the uptake of the COVID-19 vaccine, including its four subscales, by the independent characteristics. Chi-squared and Fisher exact tests (where applicable) were performed to assess the individual association of selected background characteristics with the positive attitude towards COVID-19 vaccine uptake in each of the subscales. All factors significantly (p<0.25) associated with a positive attitude towards COVID-19 vaccination at the bivariate level were thereafter included at the multivariate stage. The logistic regression was used to determine the influence of selected background characteristics on the positive attitude towards COVID-19 vaccine uptake. The adjusted odds ratios (aORs) including their 95% confidence intervals (CIs) and/or p-values are reported. Data management and analysis were conducted using Stata version 14.0 statistical software at a 5% level of significance.

### Ethical Approval

The University of Ibadan/University College Hospital Institutional Review Committee approved the survey protocol with approval number UI/EC/21/0065. Participants gave informed consent and were briefed of their freedom to withdraw from the interview at any point, before data collection. Every tenet of the Helsinki declaration and other ethical requirements were strictly complied with throughout the study. No identifying information was collected from participants and study questionnaires were accessible to only investigators and authorised research staff.

## Results

### Participants’ characteristics

The participants’ mean age was 54.3 (standard deviation [SD]: 16.3) years. Most participants were aged 50-64 years (35.7%), women (58.2%) and Yoruba (91.3%). Only about 8.0% of respondents earned less than the national minimum wage and 8.5% had no formal education. Most participants reported single health conditions (88.7%) (Table 2). Hypertension (n=116; 27.4%), diabetes mellitus (n=93; 22.0%) and heart conditions/diseases (n=83; 19.6%) were the top three conditions being managed by the patients (see Fig 1).

**Table 2.** Socio-demographic and contextual characteristics of the participants

| Characteristics | n | % |
| --- | --- | --- |
| **Age (years)** |  |  |
| <30 | 36 | 8.5 |
| 30 – 49 | 112 | 26.5 |
| 50 - 64 | 151 | 35.7 |
| ≥65 | 124 | 29.3 |
| **Sex** |  |  |
| Male | 177 | 41.8 |
| Female | 246 | 58.2 |
| **Ethnicity** |  |  |
| Yoruba | 386 | 91.3 |
| Other | 37 | 8.7 |
| **Religion** |  |  |
| Christianity | 292 | 69 |
| Islam | 131 | 31 |
| **Marital status** |  |  |
| Not married | 48 | 11.3 |
| Married | 375 | 88.7 |
| **Employment status** |  |  |
| Skilled | 313 | 74 |
| Unskilled | 110 | 26 |
| **Highest education** |  |  |
| No formal | 36 | 8.5 |
| Primary | 87 | 20.6 |
| Secondary | 130 | 30.7 |
| Higher | 170 | 40.2 |
| **Income (monthly)** |  |  |
| < ₦30,000 | 34 | 8 |
| ₦30,000 – ₦49,999 | 72 | 17 |
| ₦50,000 – ₦99,999 | 92 | 21.8 |
| ≥ ₦100,000 | 55 | 13 |
| Unknown | 170 | 40.2 |
| **Asset own** |  |  |
| Single | 307 | 72.6 |
| Multiple | 116 | 27.4 |
| **Media exposure** |  |  |
| Not exposed | 32 | 7.6 |
| Exposed | 391 | 92.4 |
| **Cooking fuel** |  |  |
| Clean | 17 | 4 |
| Unclean | 406 | 96 |
| **Drinking water source** |  |  |
| Improved | 219 | 51.8 |
| Unimproved | 204 | 48.2 |
| **Toilet facility** |  |  |
| Improved | 402 | 95 |
| Unimproved | 21 | 5 |
| **Waste disposal** |  |  |
| Hygienic | 176 | 41.6 |
| Unhygienic | 247 | 58.4 |
| **No of rooms** |  |  |
| 1 - 2 | 109 | 25.8 |
| 3 - 4 | 213 | 50.4 |
| > 4 | 101 | 23.9 |
| **No of persons** |  |  |
| 1 - 2 | 96 | 22.7 |
| 3 - 4 | 142 | 33.6 |
| 4 - 6 | 156 | 36.9 |
| > 6 | 29 | 6.9 |
| **Health condition** |  |  |
| Single | 375 | 88.7 |
| Multiple | 48 | 11.3 |

n – number of subjects per group

**Fig 1.**
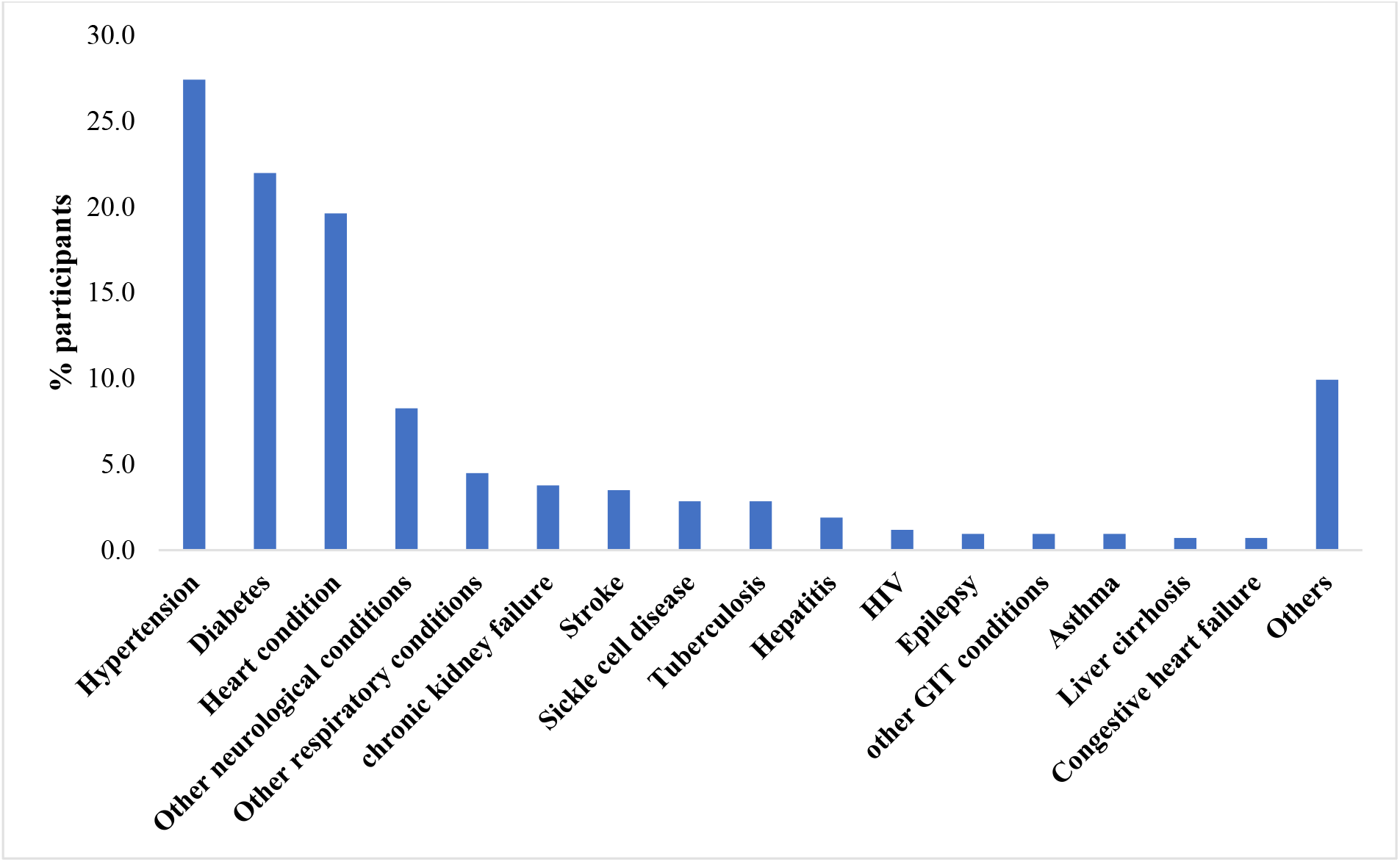
Percentages distribution of participants’ chronic health conditions (multiple response)

As shown in Table 3, majority (84.9%) of the participants rated their COVID-19 knowledge high while several (59.3%) had a poor adherence to the COVID-19 prevention protocol. Just over half (52%) had a high confidence in the government while a greater percentage had high confidence in health care workers.

**Table 3.** COVID-19 related characteristics of the participants

| Characteristics | n | % |
| --- | --- | --- |
| **COVID-19 knowledge** |  |  |
| Poor | 64 | 15.1 |
| Good | 359 | 84.9 |
| **Adherence level** |  |  |
| Poor | 251 | 59.3 |
| Better | 172 | 40.7 |
| **Full child vaccination** |  |  |
| Incomplete | 84 | 19.9 |
| Complete | 339 | 80.1 |
| **Conf. in govt** |  |  |
| Low | 143 | 33.8 |
| undecided | 60 | 14.2 |
| High | 220 | 52 |
| **Conf. in health prof** |  |  |
| Low | 42 | 9.9 |
| undecided | 44 | 10.4 |
| High | 337 | 79.7 |

n – number of subjects per group

Table 4 shows that the proportion of positive attitude towards COVID-19 vaccine uptake increased with increasing respondents’ age and levels of confidence in government handling the pandemic, in each of the subscales as well as the overall. Overall, less than half of the respondents (46.6%) had a positive attitude towards the uptake of the COVID-19 vaccine. Almost an equivalent proportion of participants had a positive attitude relating to COVID-19 vaccination against worries about unforeseen future effects (46.6%) and preferences for natural immunity (45.9%); 29.6 % against mistrust of vaccine benefit and 11.1% concerned about commercial profiteering.

**Table 4.**
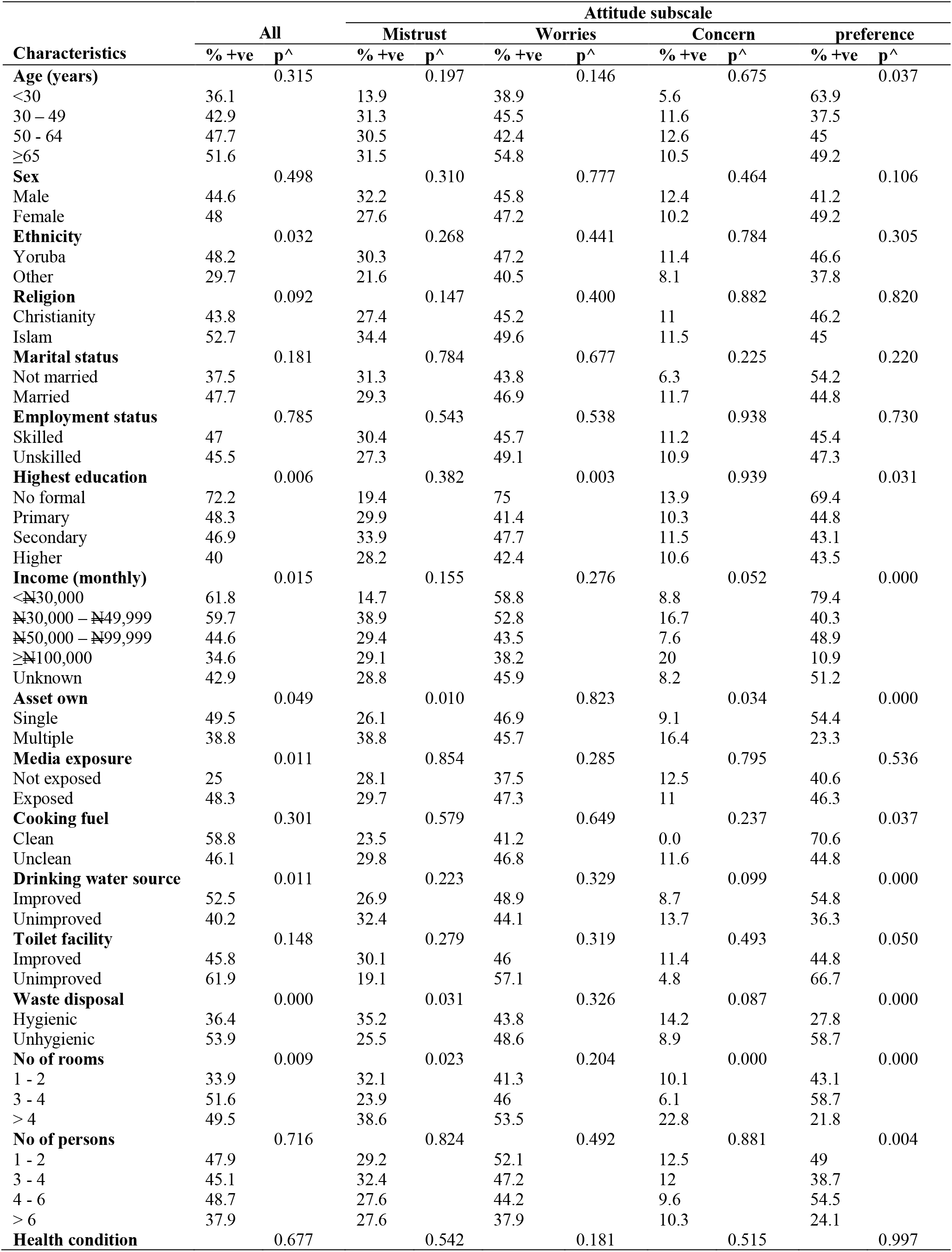
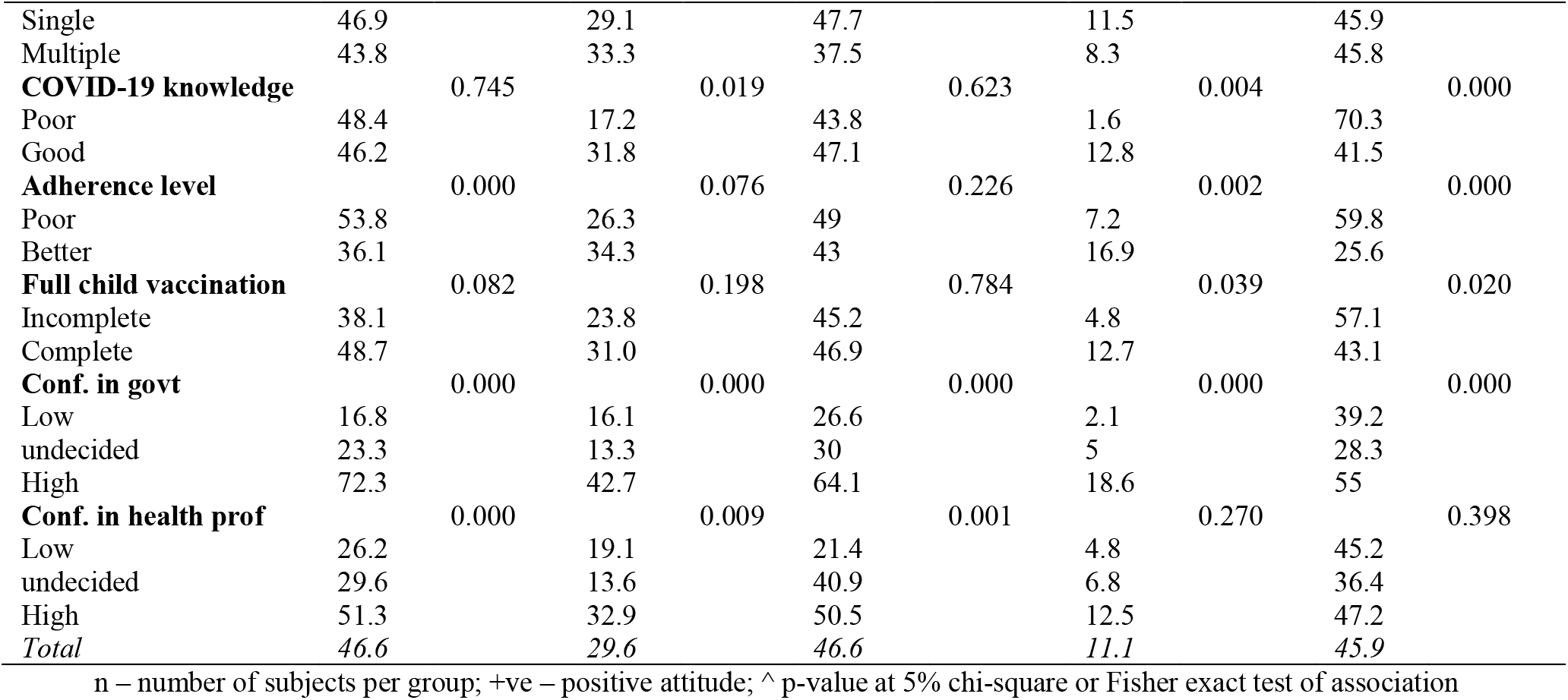
Distribution of participants and proportion of positive attitude towards COVID-19 vaccine according to selected characteristics by attitude subscales

### Factors influencing positive attitude towards COVID-19 vaccination among people living with chronic health conditions

The adjusted associations of positive attitude towards COVID-19 uptake with significant characteristics (p<0.25) at the bivariate level, including the attitude subscales, set out Model 1-4, are presented in Table 5. In the overall model, the likelihood of having a positive attitude towards COVID-19 vaccine uptake was higher among patients living in a house with more than two rooms (3-4 rooms - aOR=3.16, CI: 1.52,6.57; >4 rooms - aOR=4.29, CI: 1.87,9.87) and those who rated government high (aOR=15.78, CI:6.52,38.16) at handling the pandemic. The positive attitude was lowered among patients who had primary education (aOR=0.28; CI: 0.10,0.84) and those who had a high level of adherence to COVID-19 preventive measures (aOR=0.50; CI: 0.29,0.86). In all the models, patients who rated the government’s handling of pandemic high were more likely to have a positive attitude towards COVID-10 vaccination. Additionally, patients who owned multiple assets were more prone to having a positive attitude against mistrust of vaccine benefit (see, model 1). While patients who had formal education were less likely, those who abode in a house with many rooms were more likely to exhibit a positive attitude towards COVID-19 vaccination against worries about unforeseen future effects (see, model 2). Living in a house with many rooms significantly influenced a positive attitude against concerns about commercial profiteering (see, model 3). Patients who practised unhygienic waste disposal (aOR=2.17; CI:1.25,3.77) had higher odds of a positive attitude towards COVID-19 vaccine uptake against preferences for natural immunity. It was reduced among patients who earned more than #30,000 and among those who had better adherence to COVID-19 preventive measures (aOR=0.44; CI: 0.26,0.73) (see, model 4, Table 5).

**Table 5.** Adjusted odds ratios positive attitude towards COVID 19 vaccine uptake

| Background characteristics | Overall attitude OR (95% CI) | Attitude subscales: aOR (95% CI) |  |  |  |
| --- | --- | --- | --- | --- | --- |
|  |  | Model 1 | Model 2 | Model 3 | Model 4 |
| **Age (years)** |  |  |  |  |  |
| <30 |  | 1 | 1 |  | 1 |
| 30 – 49 |  | 2.98(0.91,9.72) | 1.28(0.55,2.97) |  | 0.45(0.13,1.61) |
| 50 - 64 |  | 2.40(0.75,7.69) | 0.83(0.36,1.92) |  | 0.65(0.18,2.38) |
| ≥65 |  | 2.34(0.70,7.77) | 1.53(0.62,3.72) |  | 0.72(0.19,2.75) |
| **Sex** |  |  |  |  |  |
| Male |  |  |  |  | 1 |
| Female |  |  |  |  | 0.99(0.58,1.67) |
| **Ethnicity** |  |  |  |  |  |
| Yoruba | 1.45(0.50,4.16) |  |  |  |  |
| Other | 1 |  |  |  |  |
| **Religion** |  |  |  |  |  |
| Christianity | 0.92(0.50,1.66) | 0.83(0.48,1.43) |  |  |  |
| Islam | 1 | 1 |  |  |  |
| **Marital status** |  |  |  |  |  |
| Not married | 1 |  |  | 1 | 1 |
| Married | 0.86(0.33,2.23) |  |  | 0.79(0.21,2.98) | 1.43(0.48,4.30) |
| **Highest education** |  |  |  |  |  |
| No formal | 1 |  | 1 |  | 1 |
| Primary | 0.28(0.10,0.84)* |  | 0.19(0.08,0.47)* |  | 0.53(0.18,1.54) |
| Secondary | 0.43(0.16,1.13) |  | 0.34(0.14,0.84)* |  | 0.61(0.21,1.77) |
| Higher | 0.58(0.19,1.77) |  | 0.30(0.12,0.74)* |  | 1.57(0.49,4.99) |
| **Income (monthly)** |  |  |  |  |  |
| <₦30,000 | 1 | 1 |  | 1 | 1 |
| ₦30,000 – ₦49,999 | 1.76(0.51,6.05) | 3.23(0.94,11.09) |  | 1.69(0.23,5.03) | 0.29(0.09,0.89)* |
| ₦50,000 – ₦99,999 | 0.53(0.14,1.98) | 1.68(0.49,5.76) |  | 0.33(0.06,1.41) | 0.29(0.09,0.94)* |
| ≥₦100,000 | 0.57(0.12,2.70) | 1.10(0.28,4.36) |  | 0.91 (0.14,4.81) | 0.06(0.01,0.29*) |
| Unknown | 0.86(0.27,2.72) | 2.10(0.63,6.98) |  | 0.52(0.10,2.16) | 0.43(0.14,1.27) |
| **Asset own** |  |  |  |  |  |
| Single | 1 | 1 |  | 1 | 1 |
| Multiple | 1.04(0.54,2.00) | 1.87(1.06,3.30)* |  | 1.43(0.60,3.36) | 0.58(0.30,1.10) |
| **Media exposure** |  |  |  |  |  |
| Not exposed | 1 |  |  |  |  |
| Exposed | 1.31(0.39,4.34) |  |  |  |  |
| **Cooking fuel** |  |  |  |  |  |
| Clean |  |  |  |  | 1.11(0.27,4.52) |
| Unclean |  |  |  |  | 1 |
| **Drinking water source** |  |  |  |  |  |
| Improved | 1 | 1 |  | 1 | 1 |
| Unimproved | 0.74(0.42,1.29) | 1.23(0.73,2.09) |  | 1.43(0.63,2.84) | 0.84(0.49,1.43) |
| **Toilet facility** |  |  |  |  |  |
| Improved | 1 |  |  |  | 1 |
| Unimproved | 2(0.46,8.71) |  |  |  | 1.08(0.23,5.01) |
| **Waste disposal** |  |  |  |  |  |
| Hygienic | 1 | 1 |  | 1 | 1 |
| Unhygienic | 1.42(0.80,2.52) | 0.63(0.36,1.12) |  | 0.82(0.38,1.80) | 2.17(1.25,3.77)* |
| **No of rooms** |  |  |  |  |  |
| 1 - 2 | 1 | 1 | 1 | 1 | 1 |
| 3 - 4 | 3.16(1.52,6.57)* | 0.75(0.40,1.41) | 1.52(0.87,2.60) | 0.64(0.24,1.69) | 1.79(0.89,3.61) |
| > 4 | 4.29(1.87,9.87)* | 1.55(0.77,3.14) | 2.38(1.29,4.40)* | 2.96(1.10,8.68)* | 0.55(0.24,1.29) |
| **No of persons** |  |  |  |  |  |
| 1 - 2 |  |  |  |  | 1 |
| 3 - 4 |  |  |  |  | 0.65(0.32,1.32) |
| 4 - 6 |  |  |  |  | 1.35(0.63,2.91) |
| > 6 |  |  |  |  | 0.73(0.22,2.41) |
| **Health condition** |  |  |  |  |  |
| Single |  |  | 1 |  |  |
| Multiple |  |  | 0.59(0.29,1.17) |  |  |
| **COVID-19 knowledge** |  |  |  |  |  |
| Poor |  | 1 |  | 1 | 1 |
| Good |  | 1.18(0.51,2.70) |  | 3.85(0.34,35.85) | 0.49(0.23,1.06) |
| **Adherence level** |  |  |  |  |  |
| Less | 1 | 1 | 1 | 1 | 1 |
| Much | 0.50(0.29,0.86)* | 1.1(0.65,1.86) | 0.84(0.54,1.31) | 1.90(0.89,3.96) | 0.44(0.26,0.73)* |
| **Full child vaccination** |  |  |  |  |  |
| Incomplete | 1 | 1 |  |  | 1 |
| Complete | 1.01(0.43,2.33) | 0.8(0.35,1.79) |  |  | 0.63(0.29,1.39) |
| **Conf. in govt** |  |  |  |  |  |
| Low | 0.80(0.33,1.95) | 1.32(0.48,3.59) | 1.21(0.57,2.60) | 0.41(0.07,2.47) | 1.29(0.55,3.04) |
| undecided | 1 | 1 | 1 | 1 | 1 |
| High | 15.78(6.52,38.16)* | 6.18(2.44,15.66)* | 5.84(2.77,12.32)* | 7.09(1.82,30.24)* | 3.6(1.58,8.19)* |
| **Conf. in health prof** |  |  |  |  |  |
| Low | 0.92(0.27,3.11) | 1.79(0.44,7.28) | 0.35(0.12,1.06) |  |  |
| undecided | 1 | 1 | 1 |  |  |
| High | 0.73(0.30,1.78) | 1.53(0.51,4.58) | 0.73(0.35,1.52) |  |  |
| -LL | 200.0 | 218.8 | 247.4 | 111.5 | 209.1 |
| N | 423 | 423 | 423 | 423 | 423 |

\*Significant at 5%; + LL - log-likelihood; N - number of observations

## Discussion

We determined COVID-19 vaccination attitude and the predictors among people living with chronic medical conditions in Ibadan, Nigeria. Less than half of the participants (46.6%) had a positive attitude towards vaccine uptake. Interestingly, the percentage reported in this study which focused solely on people with chronic conditions is very close to that reported among health workers and healthy Nigerians, [21,22]. It was however higher than the percentage reported among staff and students of a tertiary institution in south-east Nigeria, [23]. Further, the proportion reported in this study is similar to that reported among other African countries like Egypt [24] and Ghana [25]; however, it was higher than the percentage reported among Arabs [26]. In contrast, studies have shown that participants in high income countries had a higher percentage of positive attitude towards the vaccine uptake [10,27]. This could be associated with a greater trust in the vaccine because of the local production of vaccines in those countries. Another plausible reason of low percentage of positive attitudes towards the vaccine among the studied participants could be linked to the fear of side effects.

The most influential factor to having a positive attitude towards the COVID vaccine among the study participants was trust in the government’s ability to handle the pandemic. Those who trusted the government were sixteen times more likely to take the vaccine. This element of trust in the government was much stronger than the influence of confidence in health professionals. Generally, many people think, justifiably so, that the government has a lot to do in helping to manage the pandemic. Not surprising, in many other countries including Nigeria, the government was responsible for imposing curfews, ensuring COVID testing, establishing isolation centres and procuring vaccines for the people. The influence of the government had been documented before the COVID-19 pandemic as a significant predictor of flu vaccination [28]. It has also been reported to impact the COVID-19 vaccination attitude among Ireland and UK citizens [29]. This finding corroborates recent study conducted among US, Australian and UK citizens, [30]

Trust in the government and its influence on vaccination attitude does appear to be common phenomenon among people irrespective of their race. This was the case in a large intercontinental study involving twenty-six adults, [24]. Likewise, another study [31] reported the influence of mistrust of the government on vaccine attitude. There is currently no health condition that has generated so much media attention, controversy/strongly polarised opinion and political involvement as issues surrounding the COVID -19 pandemic. Hence, the citizens look up to authorities such as the government to resolve doubts and provide guidelines for the people. This view was supported by Soares et al, [32] in a study among the general Portuguese population. Park and colleagues [33] also reported the negative influence of mistrust in government on vaccination attitude among South Korean adults.

Socioeconomic status based on having more than two rooms in the house was associated with an overall positive attitude towards the vaccine. Similarly, those who owned multiple assets had a positive attitude and were more likely to trust vaccine benefits. These results are in consonance with the findings of other studies [2,10,34]. Socioeconomic status was however not associated with COVID-19 vaccination attitude among older adults in the UK, [35]. On the other hand, people of lower socioeconomic status as shown by having fewer assets, lower-income and unhygienic waste disposal, which is common among those who live in urban slums, preferred the vaccine to their natural immunity. It is possible that this group of people could not afford a good diet and felt that their immune system was not strong enough to withstand COVID infection.

Those who adhered more to the COVID preventive guidelines had a less positive attitude towards the COVID vaccine. They also preferred natural immunity to the vaccine. This could be associated with their perceived belief that they would not contract the infection by meticulously following preventive measures. Perception of COVID 19 risk has been reported to be associated with willingness to take the vaccine among health workers in Nigeria, [21]. Our results are in contrast with that of Paul and colleagues [10] who reported a high level of trust in the COVID vaccine among those with high adherence to COVID 19 measures.

The study further showed that participants who did not complete their children’s immunization showed a preponderance for natural immunity. This, however, poses a question of how far a person’s natural immunity can protect against the COVID virus. This is an aspect that requires exploration to get to the root of vaccine hesitancy. Recently, The British Society for Immunology in collaboration with the UK Coronavirus Immunology Consortium (UK-CIC) has stated that vaccination against COVID-19 is likely to lead to a more effective and longer-lasting immunity than that prompted by natural infection with the virus. The vaccine is also said to be five times more protective against the virus compared to natural immunity following the infection, [36]

Participants who rated their COVID-19 knowledge high were not likely to be concerned about commercial profiteering through the vaccine. This is very likely because many might have sought information about the infection including the vaccine and were convinced of the genuineness of the manufacturers. Hong and colleagues [37] reported the influence of COVID vaccine knowledge on vaccine acceptance.

Overall, some characteristics did not influence the vaccination attitude of participants in this study. These include gender, religion, marital status, employment status (whether skilled or unskilled). In contrast, other studies [34,38] found gender differences in vaccination attitude among French and Israelis, respectively. However, their study took place among the general population. Religious influence was seen as a strong predictor of vaccine hesitancy in a study among US adults which opined that Christian-evangelicals and black protestant were less likely to be vaccinated, [39]. Also, Jews and Arabs in Israel differed in their vaccination attitude in a study that took place in Israel, [38]. In this study, however, participants in the three main religions in Nigeria, i.e., Christianity, Islam and Traditional religions did not differ in their vaccination attitude. Again, since this study took place among people who were largely from the same Yoruba ethnic group, ethnicity rather than religion had a greater influence on vaccine attitude. In addition, as opposed to the present study, married people were more likely to receive the vaccine compared to those not married, in a study among Ugandans, [40]. Also, unlike some Indian populations where skilled and unskilled workers differed in their vaccine attitude towards commercial profiteering, [41], the reverse was the case in this study

## Conclusion

Just about half of people with chronic conditions in Ibadan Southwest Nigeria had a positive attitude towards the COVID vaccine. Their attitude was largely influenced by confidence in the government’s ability to handle the pandemic. Other mediating factors towards positive vaccine attitude were high socioeconomic status, being highly knowledgeable about COVID-19, having fully immunized children (previous vaccination attitude), poor adherence to COVID-19 preventive measures and having had no formal education.

## Recommendations/implication

The percentage of those with a positive attitude needs to be upped to achieve herd immunity. Since people with chronic conditions are more susceptible to morbidity and mortality from COVID-19 infection, a lot of attention needs to be directed at educating them on the importance of the vaccine. Different means, as well as different groups of people, must be involved in this awareness/education programme. First, the government should use communication media as well as hold physical meetings with religious and ethnic rulers to create awareness among the people. This could also increase confidence in the government.

Secondly, health care professionals need to take a more active part in both health and political issues surrounding the COVID 19 vaccines so that they do not become redundant. Thirdly, effective education aimed at improving attitude towards COVID vaccine must take cognizance of the various factors that mediate vaccination attitude. Lastly, an association of lay vaccine promoters can be formed – comprising people of different socioeconomic and cultural backgrounds. This group may help promote vaccine awareness at the grassroots level including homes, schools, marketplaces, bus terminals, among others.

## Suggestions for further study

A qualitative study on the reason for poor vaccine attitude or vaccine hesitancy.

## Data Availability

All data produced are available online at https://data.mendeley.com/datasets/nvtv8ncydh/1

## Acknowledgments

Authors are grateful to the Nursing and medical staff of the medical outpatient clinic of the University College Hospital, Ibadan for their assistance in participant recruitment.

## References

1. Kwok KO, Li KK, Wei WI, Tang A, Wong SYS, Lee SS. Influenza vaccine uptake, COVID-19 vaccination intention and vaccine hesitancy among nurses: A survey. International Journal of Nursing Studies. 2021;114.

2. Thunström L, Newbold SC, Finnoff D, Ashworth M, Shogren JF. The Benefits and Costs of Using Social Distancing to Flatten the Curve for COVID-19. Journal of Benefit-Cost Analysis. 2020;11(2):179–95.

3. Hussein NR, Naqid IA, Saleem ZSM, Almizori LA, Musa DH, Ibrahim N. A sharp increase in the number of COVID-19 cases and case fatality rates after lifting the lockdown in Kurdistan region of Iraq. Annals of Medicine and Surgery [Internet]. 2020;57(June):140–2. Available from: https://doi.org/10.1016/j.amsu.2020.07.030

4. Ohia C, Bakarey AS, Ahmad T. COVID-19 and Nigeria: putting the realities in context. International Journal of Infectious Diseases [Internet]. 2020; 95:279–81. Available from: https://doi.org/10.1016/j.ijid.2020.04.062

5. Latkin CA, Dayton L, Yi G, Konstantopoulos A, Boodram B. Trust in a COVID-19 vaccine in the U.S.: A social-ecological perspective. Social Science and Medicine. 2021;270(January).

6. Ritchie H, Mathieu E, Rodés-Guirao L, Appel C, Giattino C, Ortiz-Ospina E, et al. Coronavirus (COVID-19) Vaccinations [Internet]. 2021 [cited 2021 Feb 15]. Available from: https://ourworldindata.org/covid-vaccinations#citation

7. de Figueiredo A, Simas C, Karafillakis E, Paterson P, Larson HJ, Dariya B, et al. Mapping global trends in vaccine confidence and investigating barriers to vaccine uptake: a large-scale retrospective temporal modelling study. The Lancet [Internet]. 2020;396(10255):898–908. Available from: http://dx.doi.org/10.1016/S0140-6736(20)31558-0

8. Dubé E, Gagnon D, Ouakki M, Bettinger JA, Guay M, Halperin S, et al. Understanding vaccine hesitancy in Canada: Results of a consultation study by the Canadian Immunization Research Network. PLoS ONE. 2016;11(6):1–16.

9. Fisher KA, Bloomstone SJ, Walder J, Crawford S, Fouayzi H, Mazor KM. Attitudes Toward a Potential SARS-CoV-2 Vaccine: A Survey of U.S. Adults. Vol. 173, Annals of internal medicine. 2020. p. 964–73.

10. Paul E, Steptoe A, Fancourt D. Attitudes towards vaccines and intention to vaccinate against COVID-19: Implications for public health communications. The Lancet Regional Health -Europe. 2021; 1:100012.

11. Reiter PL, Pennell ML, Katz ML. Acceptability of a COVID-19 vaccine among adults in the United States: How many people would get vaccinated? Vol. 38, Vaccine. 2020. p. 6500–7.

12. Su Z, Wen J, Abbas J, McDonnell D, Cheshmehzangi A, Li X, et al. A race for a better understanding of COVID-19 vaccine non-adopters. Brain, Behavior, & Immunity - Health. 2020;9(September):100159.

13. OMS OM de la S. WHO SAGE values framework for the allocation and prioritization of COVID-19 vaccination. WhoCom [Internet]. 2020;(September):1–13. Available from: https://apps.who.int/iris/bitstream/handle/10665/334299/WHO-2019-nCoV-SAGE_Framework-Allocation_and_prioritization-2020.1-eng.pdf

14. Ko JY, Danielson ML, Town M, Derado G, Greenlund KJ, Kirley PD, et al. Risk Factors for Coronavirus Disease 2019 (COVID-19)–Associated Hospitalization: COVID-19– Associated Hospitalization Surveillance Network and Behavioral Risk Factor Surveillance System. Clinical Infectious Diseases. 2020;2019(Xx Xxxx):1–9.

15. Caballero AE, Ceriello A, Misra A, Aschner P, McDonnell ME, Hassanein M, et al. COVID-19 in people living with diabetes: An international consensus. Vol. 34, Journal of Diabetes and its Complications. 2020.

16. Anjorin AA. More Preparedness on Coronavirus Disease-2019 (COVID-19) in Nigeria. Pan African Journal of Life Sciences. 2020;4(1):13–6.

17. Taye GM, Bose L, Beressa TB, Tefera GM, Mosisa B, Dinsa H, et al. COVID-19 knowledge, attitudes, and prevention practices among people with hypertension and diabetes mellitus attending public health facilities in Ambo, Ethiopia. Infection and Drug Resistance. 2020; 13:4203–14.

18. Sataloff RT, Johns MM, Kost KM. DHS Comparative Reports. 2004.

19. Martin LR, Petrie KJ. Understanding the Dimensions of Anti-Vaccination Attitudes: The Vaccination Attitudes Examination (VAX) Scale. Annals of Behavioral Medicine. 2017;51(5):652–60.

20. Riccò M, Vezzosi L, Gualerzi G, Bragazzi NL, Balzarini F. Pertussis immunization in healthcare workers working in pediatric settings: Knowledge, attitudes and practices (KAP) of occupational physicians. Preliminary results from a web-based survey (2017). Vol. 61, Journal of Preventive Medicine and Hygiene. Pacini Editore S.p.A.; 2020. p. E66–75.

21. Adejumo OA, Ogundele OA, Madubuko CR, Oluwafemi RO, Okoye OC, Okonkwo KC, et al. Perceptions of the COVID-19 vaccine and willingness to receive vaccination among health workers in Nigeria. Osong Public Health and Research Perspectives. 2021;12(4):236–43.

22. Josiah B, Kantaris M. Perception of Covid-19 and Acceptance of Vaccination in Delta State Nigeria [Internet]. Vol. 21, The Nigerian Health Journal. 2021 [cited 2021 Dec 20]. p. 60–86. Available from: http://www.tnhjph.com/index.php/tnhj/article/view/510

23. Uzochukwu IC, Eleje GU, Nwankwo CH, Chukwuma GO, Uzuke CA, Uzochukwu CE, et al. COVID-19 vaccine hesitancy among staff and students in a Nigerian tertiary educational institution. Therapeutic Advances in Infectious Disease. 2021;8.

24. Mannan KA, Farhana KM. Knowledge, Attitude and Acceptance of a COVID-19 Vaccine: A Global Cross-Sectional Study. SSRN Electronic Journal. 2021;6(4).

25. Acheampong T, Akorsikumah EA, Osae-Kwapong J, Khalid M, Appiah A, Amuasi JH. Examining Vaccine Hesitancy in Sub-Saharan Africa: A Survey of the Knowledge and Attitudes among Adults to Receive COVID-19 Vaccines in Ghana. Vaccines [Internet]. 2021 Jul 22 [cited 2021 Dec 8];9(8):814. Available from: https://www.mdpi.com/2076-393X/9/8/814

26. Sallam M. COVID-19 Vaccine Hesitancy Worldwide: A Concise Systematic Review of Vaccine Acceptance Rates. Vaccines [Internet]. 2021;9(2):160. Available from: https://www.mdpi.com/2076-393X/9/2/160

27. Pogue K, Jensen JL, Stancil CK, Ferguson DG, Hughes SJ, Mello EJ, et al. Influences on attitudes regarding potential covid-19 vaccination in the United States. Vaccines. 2020;8(4):1–14.

28. Jamison A and VSFreimuthJAM, QSC, & FVS (2019). “You don’t trust a government vaccine”: N of institutional trust and influenza vaccination among AA and white adults., 221, 87–94., Quinn S, Freimuth V. Narratives of institutional trust and influenza vaccination among African American and white adults. Social Science & Medicine. 2019; 221:87–94.

29. Murphy H, Wadham C, Hassler-Hurst J. Randomized trial of a diabetes self-management education and family teamwork intervention in adolescents with Type 1 diabetes. Diabetic [Internet]. 2012 [cited 2016 Jun 2]; Available from: http://onlinelibrary.wiley.com/doi/10.1111/j.1464-5491.2012.03683.x/full

30. Trent M, Seale H, Chughtai AA, Salmon D, MacIntyre CR. Trust in government, intention to vaccinate and COVID-19 vaccine hesitancy: A comparative survey of five large cities in the United States, United Kingdom, and Australia. Vaccine. 2021.

31. Schernhammer E, Weitzer J, Laubichler MD, Birmann BM, Bertau M, Zenk L, et al. Correlates of COVID-19 vaccine hesitancy in Austria: trust and the government. Journal of Public Health. 2021;1–11.

32. Soares P, Rocha JV, Moniz M, Gama A, Laires PA, Pedro AR, et al. Factors associated with COVID-19 vaccine hesitancy. Vaccines. 2021;9(3):1–14.

33. Park HK, Ham JH, Jang DH, Lee JY, Jang WM. Political ideologies, government trust, and covid-19 vaccine hesitancy in south korea: A cross-sectional survey. International Journal of Environmental Research and Public Health. 2021;18(20).

34. Ward JK, Alleaume C, Peretti-Watel P, Seror V, Cortaredona S, Launay O, et al. The French public’s attitudes to a future COVID-19 vaccine: The politicization of a public health issue. Social Science and Medicine. 2020; 265:2016–21.

35. Williams L, Gallant AJ, Rasmussen S, Brown Nicholls LA, Cogan N, Deakin K, et al. Towards intervention development to increase the uptake of COVID-19 vaccination among those at high risk: Outlining evidence-based and theoretically informed future intervention content. British Journal of Health Psychology. 2020;25(4):1039–54.

36. Crist C. COVID Vax 5 Times More Protective Than Natural Immunity [Internet]. [cited 2021 Nov 30]. Available from: https://www.webmd.com/vaccines/covid-19-vaccine/news/20211031/covid-vax-5-times-more-protective-than-natural-immunity

37. Hong J, Xu X wan, Yang J, Zheng J, Dai S mei, Zhou J, et al. Knowledge about, attitude and acceptance towards, and predictors of intention to receive the COVID-19 vaccine among cancer patients in Eastern China: A cross-sectional survey. Journal of Integrative Medicine [Internet]. 2021;(xxxx). Available from: https://doi.org/10.1016/j.joim.2021.10.004

38. Green MS, Abdullah R, Vered S, Nitzan D. A study of ethnic, gender and educational differences in attitudes toward COVID-19 vaccines in Israel – implications for vaccination implementation policies. Israel Journal of Health Policy Research. 2021;10(1):1–12.

39. Whitehead AL, Perry SL. How Culture Wars Delay Herd Immunity: Christian Nationalism and Anti-vaccine Attitudes. Socius. 2020;6.

40. Echoru I, Ajambo PD, Keirania E, Bukenya EEM. Sociodemographic factors associated with acceptance of COVID-19 vaccine and clinical trials in Uganda: a cross-sectional study in western Uganda. BMC Public Health. 2021;21(1):1–8.

41. Godasi GR, Donthu RK, Mohammed AS, Pasam RS, Tiruveedhula SL. Attitude towards COVID-19 vaccine among the general public in south India: A cross sectional study. Archives of Mental Health. 2021 Jan 1;22(1):28–35.

